# Polygenic Risk for Depression and Anterior and Posterior Hippocampal Volume in Children and Adolescents

**DOI:** 10.1101/2023.05.11.23289784

**Authors:** Hailee Hurtado, Melissa Hansen, Jordan Strack, Uku Vainik, Alexandra L. Decker, Budhachandra Khundrakpam, Katherine Duncan, Amy S. Finn, Donald J. Mabbott, Emily C. Merz

## Abstract

**Background:** Depression has frequently been associated with smaller hippocampal volume. The hippocampus varies in function along its anterior-posterior axis, with the anterior hippocampus more strongly associated with stress and emotion processing. The goals of this study were to examine the associations among parental history of anxiety/depression, polygenic risk scores for depression (PGS-DEP), and anterior and posterior hippocampal volumes in children and adolescents. To examine specificity to PGS-DEP, we examined associations of educational attainment polygenic scores (PGS-EA) with anterior and posterior hippocampal volume.

**Methods:** Participants were 350 3– to 21-year-olds (46% female). PGS-DEP and PGS-EA were computed based on recent, large-scale genome-wide association studies. High-resolution, T1-weighted magnetic resonance imaging (MRI) data were acquired, and a semi-automated approach was used to segment the hippocampus into anterior and posterior subregions.

**Results:** Children and adolescents with higher PGS-DEP were more likely to have a parent with a history of anxiety/depression. Higher PGS-DEP was significantly associated with smaller anterior but not posterior hippocampal volume. PGS-EA were not associated with anterior or posterior hippocampal volumes.

**Limitations:** Participants in these analyses were all of European ancestry.

**Conclusions:** Polygenic risk for depression may lead to smaller anterior but not posterior hippocampal volume in children and adolescents, and there may be specificity of these effects to PGS-DEP rather than PGS-EA. These findings may inform the earlier identification of those in need of support and in the future could inform the design of more effective, personalized treatment strategies.

## Introduction

Depression is prevalent and a leading cause of disability worldwide, making it a major public health issue (Herrman et al., 2022). The core symptoms of depressive disorders are depressed mood and anhedonia. Although depression is relatively rare in childhood, it increases in prevalence starting in adolescence (Avenevoli et al., 2015; Kessler et al., 2005). The etiology of depression is multifactorial and includes a robust genetic component (Flint & Kendler, 2014). At the neural level, depression has been repeatedly associated with smaller volume in the hippocampus (Belleau et al., 2019; Schmaal et al., 2020), which is crucial to learning, memory, and emotion processes and highly susceptible to chronic stress (Kaul et al., 2021; McEwen et al., 2016). Genome-wide polygenic scores are derived from genome-wide association studies (GWAS) and have been used to elucidate genetic influences on a range of phenotypes (Armstrong-Carter et al., 2021). Polygenic risk scores for depression (PGS-DEP), computed via weighted sums of the single nucleotide polymorphisms (SNPs) associated with depression, have been found to predict depressive symptoms and risk for depression in multiple studies (Howard et al., 2019; Wray et al., 2018). Yet, the associations between PGS-DEP and hippocampal structure are not well understood. As such, one main goal of this study was to examine the associations between PGS-DEP and hippocampal structure in children and adolescents.

### PGS-DEP and Depression

Several large-scale GWAS of depression have been conducted (Howard et al., 2019; Wray et al., 2018). One of the largest and most recent included meta-analyzed data on 807,553 individuals and identified 102 independent genetic variants associated with depression (Howard et al., 2019). PGS-DEP derived from these GWAS significantly predict depressive symptoms and risk for depression in independent samples of adults (Mitchell et al., 2021) and children and adolescents (Halldorsdottir et al., 2019; Kwong et al., 2021; Rice et al., 2019). An important next step is to build an understanding of how PGS-DEP influences brain structure and function, leading to variability in mental health outcomes.

### Anterior and Posterior Hippocampal Function

The hippocampus varies in function along its anterior-posterior axis (Grady, 2020; Poppenk et al., 2013; Strange et al., 2014). In rodent studies, the dorsal subregion (posterior in humans) shows stronger associations with spatial learning and memory, while the ventral subregion (anterior in humans) shows stronger associations with emotion processing (Bannerman et al., 2003, 2004; Fanselow & Dong, 2010; Kjelstrup et al., 2002; Levone et al., 2021; Maren & Holt, 2004; Trivedi & Coover, 2004). Human functional neuroimaging studies have yielded findings consistent with these results (Kumaran et al., 2009; Nadel et al., 2013; Poppenk & Moscovitch, 2011). These distinct roles are thought to be due to differential connectivity patterns, with the posterior hippocampus strongly connected to regions such as the retrosplenial cortex, cuneus, precuneus, cingulate cortex, and inferior parietal cortex (Dalton et al., 2019; Poppenk & Moscovitch, 2011; Tang et al., 2020) and the anterior hippocampus strongly connected to emotion processing regions (e.g., amygdala, medial prefrontal cortex) (Adnan et al., 2016; Blessing et al., 2016; Satpute et al., 2012).

### Depression and Anterior and Posterior Hippocampal Structure

While major depressive disorder (MDD) is associated with volumetric reductions in both anterior and posterior hippocampal subregions (Belleau et al., 2019; Liu et al., 2021; Malykhin et al., 2010), there is evidence that the anterior hippocampus might be particularly vulnerable. In animal studies, chronic stress, which often precedes depression, has stronger effects on the ventral compared to dorsal hippocampus (Hawley et al., 2012; Hawley & Leasure, 2012).

Moreover, both depression and antidepressants have stronger effects on ventral compared to dorsal hippocampal morphology (O’Leary & Cryan, 2014; Tanti & Belzung, 2013; Willard et al., 2009). Specifically, chronic stress reduces neurogenesis in the ventral hippocampus and antidepressants reverse these effects (Levone et al., 2015). In humans, treatment for depression (electroconvulsive therapy) has been associated with increases in anterior hippocampal volume (Gyger et al., 2021; Joshi et al., 2016). Taken together, this work in both humans and animals raises the possibility that higher PGS-DEP may lead to volumetric reductions that are stronger in the anterior compared to the posterior hippocampus.

### PGS-DEP and Hippocampal Structure

Despite the link between depression and smaller hippocampal volumes, large-scale studies have not found that PGS-DEP are associated with total hippocampal volumes in adults (Reus et al., 2017) or children (Alemany et al., 2019). These non-significant findings may reflect variability in the strength of associations by hippocampal subregion. To our knowledge, only one study has examined the associations between PGS-DEP and hippocampal subregional volumes. In this large-scale study, PGS-DEP were unrelated to hippocampal head, body, or tail volumes in children (Pine et al., 2023). The hippocampal head corresponds to the anterior hippocampus, while hippocampal body and tail are considered parts of the posterior hippocampus (Botdorf et al., 2022). However, Pine et al. (2023) was restricted to 9– to 11-year-olds and fully automatically segmented the hippocampus. Importantly, in contrast to Pine et al. (2023), we used a manual segmentation approach to subdivide the automatically segmented hippocampal labels into anterior and posterior portions (Decker et al., 2020). In addition, our sample included adolescents, an age range in which the effects of PGS-DEP might be more likely to manifest compared to childhood (Avenevoli et al., 2015).

### Parental History of Depression and PGS-DEP in Children

Parental history of depression heightens risk for depression in children and adolescents (Pagliaccio et al., 2020; Weissman et al., 2016), likely because of a combination of genetic and environmental mechanisms. Parental history of depression has also been associated with smaller hippocampal volume in children and adolescents, although findings have been inconsistent (Kemp et al., 2022; Nazarova et al., 2022), with some studies showing significant associations (Chen et al., 2010; Rao et al., 2010) and others showing no associations (Lupien et al., 2011; Mannie et al., 2014; Pagliaccio et al., 2020). In one study, maternal history of depression was associated with reduced bilateral hippocampal head volume and increased left hippocampal body volume in children (Hubachek et al., 2021). Parents contribute genetics to their children while also shaping the rearing environments their children experience. Parents with depression may pass down a genetic predisposition for depression to their children and provide more stressful rearing environments to their children compared to parents without depression (Goodman & Gotlib, 1999). Yet, less is understood about whether polygenic risk for depression represents a pathway through which parental history of depression may lead to reduced hippocampal volume in children.

In sum, the current study addresses novel questions about associations of PGS-DEP with anterior and posterior hippocampal volume in a non-clinical sample of children and adolescents. It is critical to understand these associations in young individuals to shed light on how these effects unfold prior to the onset or worsening of depression. Understanding the early emergence of these associations could support the earlier identification of those at risk for depression and guide decision-making that leads to more effective prevention and intervention strategies.

### Current Study

The main goals of this study were to investigate the associations between PGS-DEP and anterior and posterior hippocampal volume and whether PGS-DEP mediated the association of parental history of anxiety/depression with anterior or posterior hippocampal volume in children and adolescents. We also examined the associations of educational attainment polygenic scores (PGS-EA) with anterior and posterior hippocampal volume to discern whether patterns of results found for PGS-DEP were specific to PGS-DEP rather than PGS-EA. Participants were 3– to 21– year-olds (*M* = 12.07 years; *N* = 350; 46% female). High-resolution, T1-weighted MRI data were acquired, and anterior and posterior hippocampal subregions were segmented using a semi-automated approach (Decker et al., 2020). PGS-DEP and PGS-EA were computed using summary statistics from recent, well-powered GWAS of depression (Howard et al., 2019) and educational attainment (Lee et al., 2018), respectively. We hypothesized that PGS-DEP would be more strongly associated with anterior compared to posterior hippocampal volume. We also expected that anterior hippocampal volume would be more strongly associated with PGS-DEP compared to PGS-EA. PGS-DEP-by-age interactions were examined for hippocampal subregional volumes because the effects of PGS-DEP on hippocampal structure may be stronger in older adolescents who have already gone through puberty (Avenevoli et al., 2015).

## Methods

### Participants

This study utilized data from the Pediatric Imaging, Neurocognition, and Genetics (PING) study, which enrolled typically-developing children and adolescents; the PING data set is publicly available (Jernigan et al., 2016). For this study, participants were recruited across nine sites in the United States. Individuals with neurological disorders; history of head trauma; preterm birth (< 36 gestational weeks); diagnosis of autism spectrum disorder, bipolar disorder, schizophrenia, or intellectual disability; pregnancy; daily illicit drug use by the mother for more than one trimester; or contraindications for MRI were excluded from the study (Jernigan et al., 2016). More common forms of psychopathology such as anxiety, depression, and attention-deficit/hyperactivity disorder were not excluded from the PING study because the recruitment strategy was designed to be representative of the general population (Newman et al., 2015). Participants in this investigation ranged from 3 to 21 years of age, and 46% were female (see Table 1 for sample characteristics and Figure S1 for a histogram showing the age distribution).

**Table 1.** Descriptive statistics for sample characteristics and anterior and posterior hippocampal volume (*N* = 350)

|  | *M* | *SD* | **Range** |
| --- | --- | --- | --- |
| Age (years) | 12.07 | 4.71 | 3.58-21.00 |
| Family income (U.S. dollars) | 117,980.00 | 74,515.54 | 4,500.00-325,000.00 |
| Parental education (years) | 15.62 | 1.90 | 8.00-18.00 |
| Anterior hippocampal volume (mm<sup>3</sup>) | 2425.84 | 323.59 | 1645.52-3337.18 |
| Posterior hippocampal volume (mm<sup>3</sup>) | 2322.83 | 276.18 | 1504.27-3175.46 |
|  | *%* | *n* | **Range** |
| Sex (female) | 46.29 | 162 | -- |
| Parental history of anxiety/depression | 29.14 | 102 | -- |

*Note.* The data for anterior and posterior hippocampal volume are adjusted for intracranial volume (ICV). U.S., United States; --, not applicable

Written informed consent was provided by parents for all participants younger than 18 years of age and by the participants themselves if they were 18 years or older. Child assent was obtained for 7– to 17-year-old participants. Each site’s Institutional Review Board approved the study.

### Genomic Data

The PING dataset includes 550,000 SNPs genotyped from saliva samples using Illumina Human660W-Quad BeadChip. Computation of polygenic scores followed steps similar to that of our previous studies (Khundrakpam et al., 2020; Merz et al., 2022). Steps included preparation of the data for imputation using the “imputePrepSanger” pipeline (https://hub.docker.com/r/eauforest/imputeprepsanger/) and implemented on CBRAIN (Sherif et al., 2014) using Human660W-Quad_v1_A-b37-strand chip as reference. The next step involved data imputation with Sanger Imputation Service (McCarthy et al., 2016) using default settings and the Haplotype Reference Consortium (HRC) (http://www.haplotype-reference-consortium.org/) as the reference panel. Using Plink 1.9 (Chang et al., 2015), the imputed SNPs were then filtered with the inclusion criteria: SNPs with unique names, only ACTG, and minor allele frequencies (MAF) > .05. All SNPs that were included had INFO scores R^2^ > .9 with Plink 2.0. Next, using PRSice 2.1.2 (Euesden et al., 2015) additional ambiguous variants were excluded, resulting in 4,696,385 variants being available for polygenic scoring. We filtered individuals with .95 loadings to the European principal component (GAF_Europe variable provided with the PING data), resulting in 526 participants. These participants were then used to compute 10 principal components with Plink 1.9. PGS-DEP computed based on Howard et al (2019), and PGS-EA based on the EA3 GWAS (Lee et al., 2018) were used in analyses. We clumped the data as per PRSice default settings (clumping distance = 250 kb, threshold r^2^ = 0.1). PGS-DEP calculated at nine *p*-value thresholds (1 x 10^-6^, 1 x 10^-5^, .0001, .001, .01, .05, .1, .5, and 1), which correspond to the level of significance needed for SNPs to be included in the polygenic score, were used in this study. The number of SNPs included at each of these *p*-value thresholds is provided in Table S1.

### Image Acquisition and Processing

Each PING study site used a standardized structural MRI protocol, with data collected using 3-Tesla scanners manufactured by General Electric, Siemens, and Philips (Jernigan et al., 2016). T1-weighted images were acquired using a high-resolution 3D RF-spoiled gradient echo sequence (Jernigan et al., 2016; Merz et al., 2022). Details of the imaging acquisition and processing can be found in Jernigan et al. (2016). The raw T1-weighted imaging data for the PING study are publicly available for a subset of the sample (https://nda.nih.gov). These data were used in this study.

Hippocampal volumetric segmentation was conducted for a previous study (Decker et al., (2020), which provides a full description of these methods. Prior to segmenting the hippocampus, we visually inspected the images to check for indicators of motion using Display (version 2.0), an MRI image viewing software (http://www.bic.mni.mcgill.ca/software/Display/Display.html). Images were rated based on the degree to which signs of motion were detectable on a scale ranging from 1 (no signs of motion) to 4 (excessive motion). Images that had either clear or excessive signs of motion were excluded from analyses (Decker et al., 2020).

### Hippocampal Segmentations

We used a combination of automatic and manual methods to define the anterior and posterior hippocampus. First, we segmented the whole hippocampus automatically using the Multiple-Automatically Generated Templates for different brains algorithm (MAGeT Brain) (Pipitone et al., 2014), which has been validated in clinical and healthy adult samples and generates labels for the whole hippocampus that are comparable to existing automated methods (Pipitone et al., 2014). The MAGeT Brain algorithm uses a set of manually labeled hippocampal atlases as inputs to segment unlabeled T1 images in a dataset. We used five pre-existing, manually segmented hippocampal atlases (Winterburn atlases) that included definitions of hippocampal subfields as inputs (Winterburn et al., 2013), which have previously been used in analyses validating MAGeT Brain (Pipitone et al., 2014) and span the length of the anterior-posterior hippocampal axis. The MAGeT Brain algorithm registers these manually labeled atlas labels via nonlinear image registration to a subset of MR images in the sample specified as template images. Each of the newly generated labels on each template image is then registered to the entire dataset of MR images. The labels on each MR image are then fused using a voxel voting procedure, in which the most commonly occurring label at each voxel is retained as part of the final label. By registering the atlases to a subset of the sample, the templates, and then using the template labels to segment the entire dataset, labeling errors that might arise due to anatomical differences between the atlases and subject images are minimized (Pipitone et al., 2014). Hippocampal atlases developed by the same group that are based on adult anatomy and used in conjunction with MAGeT (Pipitone et al., 2014) have been validated in a developmental sample (Guo et al., 2015). This study showed that MAGeT in conjunction with adult hippocampal atlases produce accurate labels in a developmental sample relative to manually derived labels (Guo et al., 2015). This suggests that using adult atlases to segment the developing hippocampus with the MAGeT algorithm results in labels that have acceptable accuracy relative to manually derived labels.

After automated segmentation of the whole hippocampus, we combined the subfield labels (CA1, CA2-3, CA4-DG, subiculum, SRSLSM) into a single label for the left and right hippocampus. We then visually inspected each label to ensure that the label covered the hippocampus. Data were included if the segmentations covered the entire hippocampus on each slice that the hippocampus was visible or most of the hippocampus on each slice that the hippocampus was visible. Otherwise, data were excluded. Two labels for which the segmentations only covered a few (3 or 4) slices of hippocampus were excluded.

A trained rater (A.L.D.) then identified the slice that subdivided the anterior and posterior segments on the remaining images by identifying the slice corresponding to the uncal apex, which is a commonly used landmark for the anterior-posterior hippocampus boundary (Weiss et al., 2005). The caudal-most slice of the anterior hippocampus corresponded to the last slice at which the uncal apex was visible, and the rostral most slice of the posterior hippocampus corresponded to the first slice at which the uncal apex was no longer visible. To ensure the accuracy of this boundary, a second trained rater re-identified the anterior-posterior boundary in an overlapping 10% of the labels. For the anterior-posterior boundary, the researchers identified the same slice in 94% of cases, and the same or a slice that differed by one slice in 98% of cases.

Both raters were blind to demographic information relevant to this study (age, sex, family income). After identifying the boundary slice, any part of a subfield label (CA1, CA2/3, CA4/DG, subiculum, SRLM) rostral to the uncus was counted towards the volume of the anterior hippocampus, whereas any part of a label that was caudal to the uncus was counted towards the volume of the posterior hippocampus. Thus, the volume of the anterior and posterior segments, respectively, reflected the total voxels covered by any subfield label that was rostral or caudal to the boundary slice. In this way, the subfield labels were largely treated as though they were a single label: we ignored the divisions between them, and they were not used to inform the boundary between anterior and posterior segments. Data for anterior and posterior hippocampal volume were then adjusted for intracranial volume (ICV) using a regression-based approach (Jack et al., 1989) described by Decker et al. (2020) (see also Supplemental Materials). These adjusted volumetric data were used in analyses.

Of the 526 participants with polygenic score data, 350 had anterior and posterior hippocampal volume data, and 522 had parental history of anxiety/depression data. Thus, 350 participants were included in analyses of associations between the polygenic scores and anterior and posterior hippocampal volume, and 522 were included in analyses of associations between parental history of anxiety/depression and PGS-DEP.

### Parental History of Anxiety/Depression

Respondents indicated whether the biological mother and/or father of the participant had a history of anxiety or depression. These responses were summed to create a measure of the number of parents with a known history of anxiety or depression (0, 1, or 2). A dichotomous variable was then created in which 0 = no parental history of anxiety/depression and 1 = one or two parents with a history of anxiety/depression.

### Statistical Analyses

Multiple linear regression analyses in SAS (version 9.4) were conducted using the general linear model procedure to examine associations of PGS-DEP and PGS-EA (at nine different *p*-value thresholds) with anterior and posterior hippocampal volume. False discovery rate (FDR) corrections were employed to control for multiple comparisons (Benjamini & Hochberg, 1995). Covariates included age, sex, family income, and scanner/site. In the PING study, 12 MRI scanners were used across the nine data collection sites; therefore, scanner rather than site was included as a covariate in analyses of hippocampal volume. Age-squared was initially included as a covariate in analyses of hippocampal volume but was not included in the final models because it was not significant. The first 10 principal components (PC1-10) were also included as covariates in all analyses involving PGS-DEP or PGS-EA (Price et al., 2006). Including PC1-10 in analyses controls for random differences in population genomic signatures that may explain outcomes (Price et al., 2006). Effect sizes (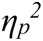) are presented, with values of .01, .06, and .14 indicating small, medium, and large effects, respectively (Cohen, 1988). We also tested whether the correlation between PGS-DEP and anterior hippocampal volume was statistically different than the correlation between PGS-DEP and posterior hippocampal volume. We used Meng, Rosenthal, and Rubin’s z test (Meng et al., 1992) via the cocor package in R (Diedenhofen & Musch, 2015).

We then examined whether PGS-DEP mediated the association between parental history of anxiety/depression and anterior and/or posterior hippocampal volume in children and adolescents. First, we examined associations between parental history of anxiety/depression and PGS-DEP while controlling for age, sex, PC1-10, and site. If these associations were significant and significant associations were found between PGS-DEP and anterior or posterior hippocampal volume (as described above), we then tested the significance of the indirect effect (*ab* path) using bias-corrected bootstrapping via the PROCESS macro in SAS (Hayes, 2013; MacKinnon et al., 2002). Indirect effects were significant if the 95% confidence intervals (CIs) did not include zero (Preacher & Hayes, 2008).

## Results

Descriptive statistics for anterior and posterior hippocampal volume are provided in Table 1, and zero-order correlations between PGS-DEP, parental history of anxiety/depression, and anterior and posterior hippocampal volume are provided in Table S2. As shown in Table S2, higher PGS-DEP was significantly correlated with a parental history of anxiety/depression and smaller anterior hippocampal volume.

### Associations between PGS-DEP and Anterior and Posterior Hippocampal Volume

Higher PGS-DEP at *p*-value thresholds of .10, .05, and .001 were significantly associated with reduced anterior hippocampal volume (see Table 2 and Figure 1). The effect size (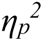) was .02 for PGS-DEP computed at all three *p*-value thresholds. PGS-DEP was not significantly associated with posterior hippocampal volume. The correlation between PGS-DEP and anterior hippocampal volume was significantly different than that between PGS-DEP and posterior hippocampal volume for PGS-DEP at *p*-value thresholds of .10 (*z* = –2.13, *p* = .03) and .05 (*z* = – 2.02, *p* = .04) but not for PGS-DEP at a *p*-value threshold of .001 (*z* = –1.49, *p* = .14). There were no significant PGS-DEP-by-age interactions for anterior or posterior hippocampal volume.

**Table 2.**
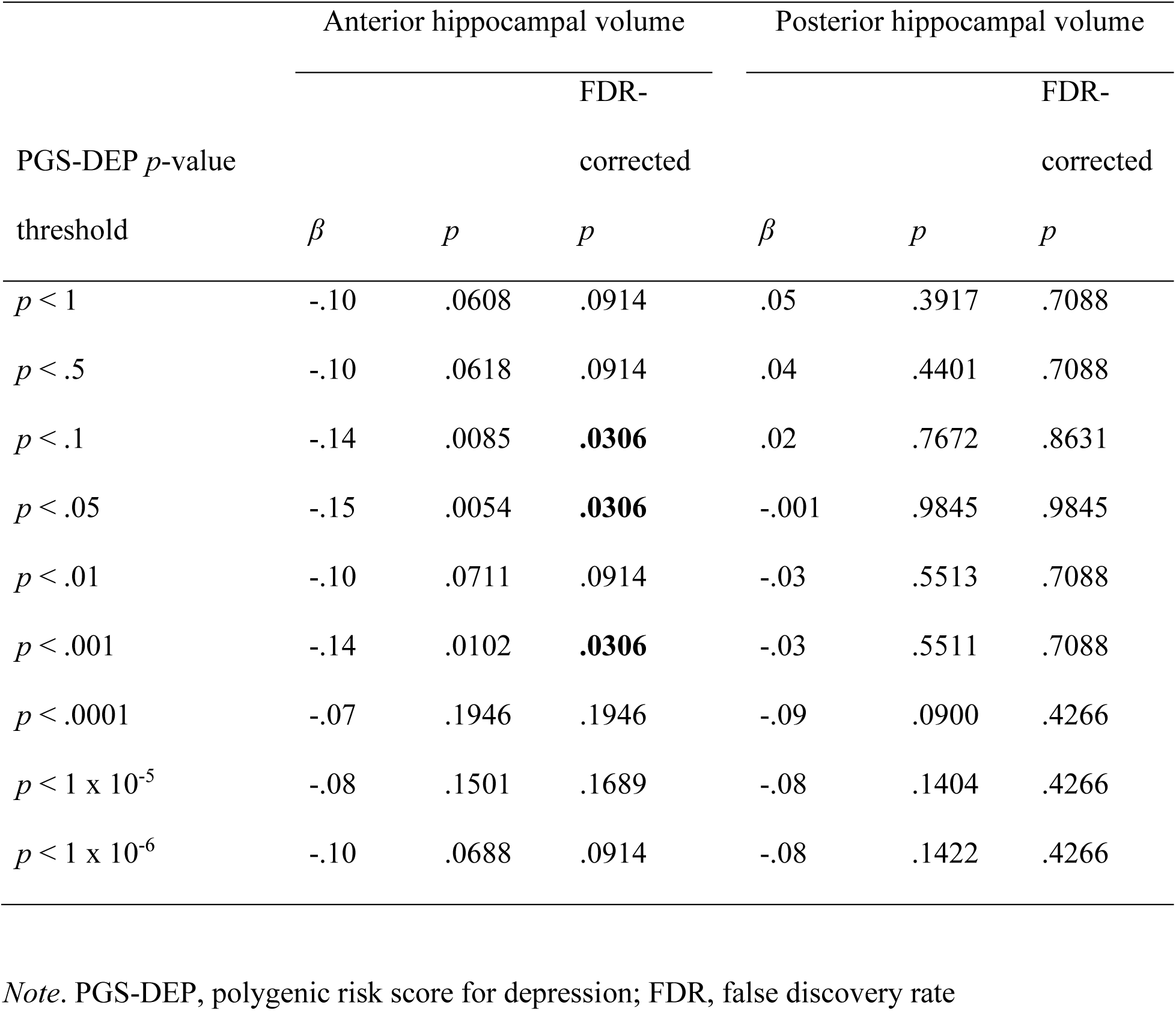
Multiple linear regression results showing associations between PGS-DEP and anterior and posterior hippocampal volume

**Figure 1.**
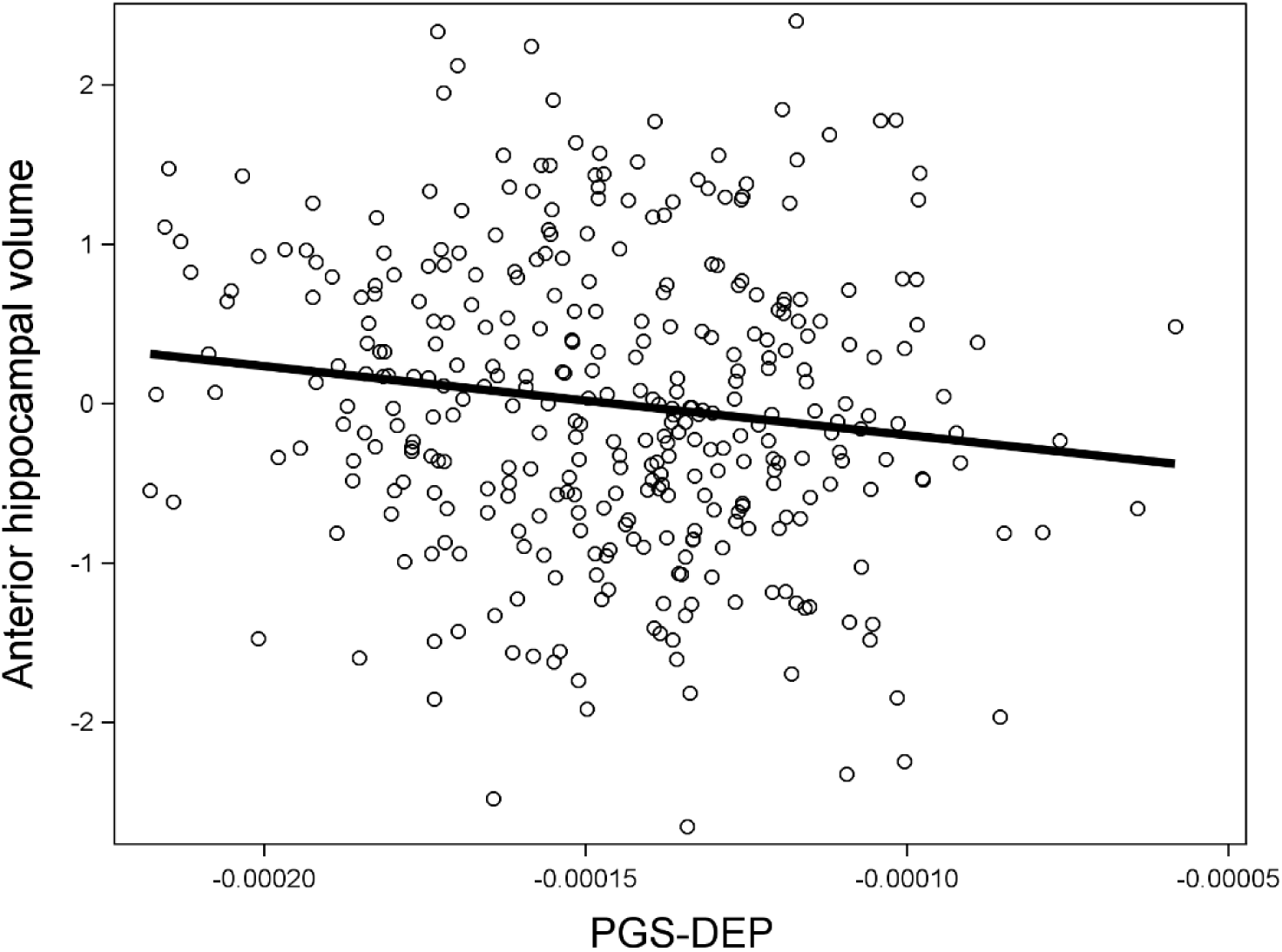
Higher polygenic risk for depression (PGS-DEP) was significantly associated with smaller anterior hippocampal volume. Data for PGS-DEP computed at a *p*-value threshold of .10 are displayed. Anterior hippocampal volume data were adjusted for covariates.

### Associations between PGS-EA and Anterior and Posterior Hippocampal Volume

There were no significant associations between PGS-EA and anterior or posterior hippocampal volume (see Table S3).

### Parental History of Anxiety/Depression, PGS-DEP, and Anterior Hippocampal Volume

Having a parent with a history of anxiety/depression was associated with higher PGS-DEP at *p*-value thresholds of .05 (*β* = .22, *p* = .03, 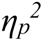 *=* .01) and .10 (*β* = .24, *p* = .02, 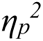 = .01) but not at a *p*-value threshold of .001 (*β* = .15, *p* = .14). In addition, there was a significant indirect effect of parental history of anxiety/depression on anterior hippocampal volume via PGS-DEP, *ab* = –.03, 95% CI: –.0949, –.0004. Having a parent with a history of anxiety/depression was associated with higher PGS-DEP which was in turn associated with smaller anterior hippocampal volume (see Figure 2).

**Figure 2.**
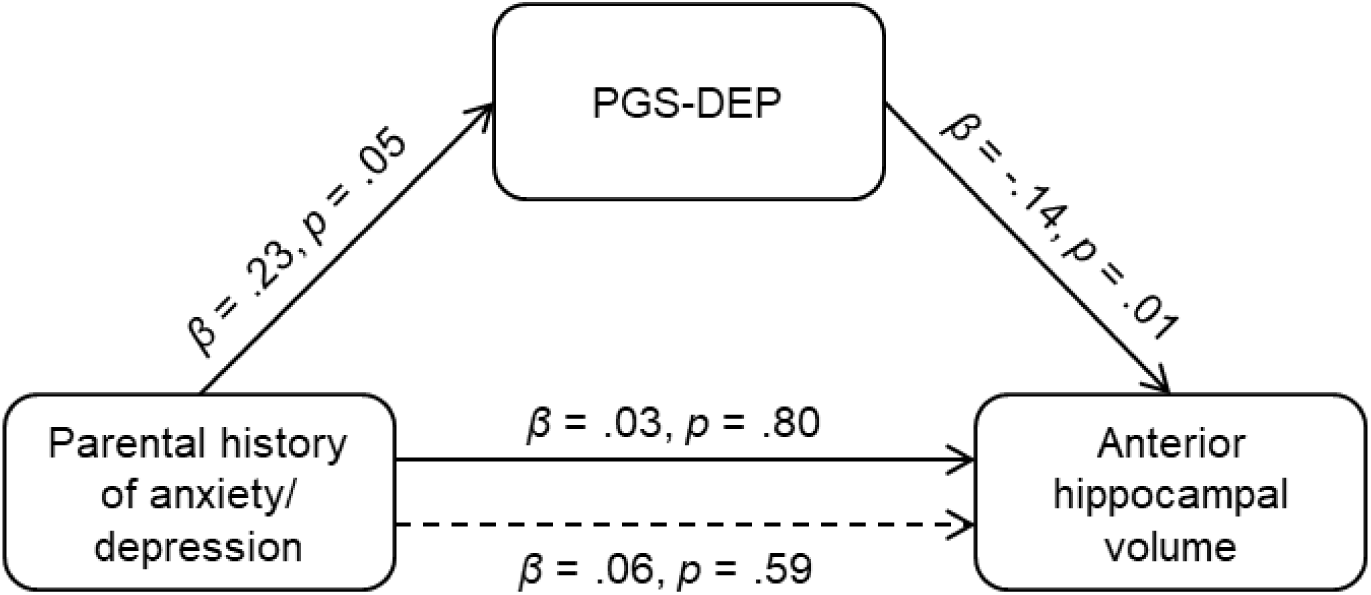
Significant indirect effect of parental history of anxiety/depression on anterior hippocampal volume via polygenic risk for depression (PGS-DEP; *p*-value threshold: .10), *ab* = –.03, 95% CI: –.0949, –.0004. Having a parent with a history of anxiety/depression was associated with higher polygenic risk for depression, which was associated with smaller anterior hippocampal volume in children and adolescents.

## Discussion

Here, we investigated the associations between PGS-DEP and anterior and posterior hippocampal volume in a large sample of children and adolescents. We also examined whether PGS-DEP were associated with a greater likelihood of having a parental history of anxiety/depression. Higher PGS-DEP was significantly associated with reduced anterior but not posterior hippocampal volume, consistent with the idea that anterior hippocampal volumes are particularly vulnerable in individuals who are at higher genetic risk for depression. There was also a significant indirect association between parental history of anxiety/depression and smaller anterior hippocampal volume via higher PGS-DEP. Having a parent with a history of anxiety/depression was associated with higher PGS-DEP, which was in turn associated with smaller anterior hippocampal volume.

### Higher PGS-DEP is Associated with Smaller Anterior Hippocampal Volume

The few previous studies that have investigated associations between PGS-DEP and total hippocampal volume have yielded non-significant findings (Alemany et al., 2019; Reus et al., 2017). And, one previous study did not find significant associations between PGS-DEP and hippocampal head, body, or tail volumes in children (Pine et al., 2023). The difference between our results and those of Pine et al. (2023) could be partially due to differences in sample characteristics. More specifically, Pine et al. (2023) focused on 9– to 11-year-olds, whereas our sample included adolescents. In addition, we used a manual segmentation approach to subdivide automatically generated hippocampal labels into anterior and posterior portions (Decker et al., 2020).

Higher genetic risk for depression may lead to reductions in anterior hippocampal volume that then contribute to increased risk for depression. Both animal models and human studies have indicated that the anterior hippocampus is heavily involved in emotion processing (Adnan et al., 2016; Blessing et al., 2016; Fanselow & Dong, 2010) likely due in part to functional connections with regions such as the amygdala and ventromedial prefrontal cortex (Adnan et al., 2016; Blankenship et al., 2017; Poppenk & Moscovitch, 2011). Genetic effects on anterior hippocampal morphology may lead to altered emotional memory and regulation in ways that increase risk for depression. In addition, altered anterior hippocampal morphology may lead to reduced negative feedback control over the hypothalamic-pituitary-adrenal (HPA) axis, leading to a disinhibited stress response which in turn increases risk for depression (Blessing et al., 2016; Frodl & O’Keane, 2013). In this way, genetic effects leading to reduced anterior hippocampal morphology may contribute to increases in vulnerability to chronic stress (Anacker et al., 2018; Jones et al., 2022; Levone et al., 2015). Given that the anterior hippocampus likely plays many roles, there may be additional implications of volumetric changes in this structure for emotion processing and aspects of memory functioning (Strange et al., 2014).

While findings from previous studies are consistent with the notion that smaller hippocampal volume may precede the onset of depression (Chen et al., 2010; Rao et al., 2010), in studies of MDD, recurrent major depressive episodes and prolonged duration of the disorder have also been associated with reductions in hippocampal volume (Belleau et al., 2019; Schmaal et al., 2020). The PING sample is a non-clinical sample of children and adolescents. However, participants with depression were not excluded, and depressive symptoms were not measured for the full sample. Thus, although many participants likely did not have a clinically significant depressive disorder, some probably did (Avenevoli et al., 2015; Kessler et al., 2005). Smaller hippocampal volume may be both a cause and consequence of depression. Patterns of altered hippocampal subregional volumes may be different in high-risk individuals prior to the onset of depression compared to after recurrent depressive episodes. Reductions in anterior hippocampal volume may emerge prior to more widespread reductions across the longitudinal axis of the hippocampus (Hubachek et al., 2021).

Effect sizes were small; after taking the covariates into account, PGS-DEP uniquely explained 2% of the variability in anterior hippocampal volume. Although genetic risk, chronic stress, and possibly reduced anterior hippocampal volume are risk factors for depression, it is how these risk factors relate to one another (e.g., diathesis-stress model) and accumulate that explains the most variability in mental health outcomes (Belleau et al., 2019; McEwen et al., 2016). Future longitudinal MRI studies of PGS-DEP are needed that measure chronic stress, anterior and posterior hippocampal volumes, and depressive symptoms over time to elucidate how effects of these variables on each other unfold over time.

In contrast to the results for PGS-DEP, PGS-EA was not associated with either anterior or posterior hippocampal volumes. Previous studies have distinguished between polygenic scores for psychiatric disorders and those for cognitive outcomes, such as educational attainment (Alemany et al., 2019; Kwong et al., 2021). Our results suggest that smaller anterior hippocampal volume may be more strongly associated with genetic factors associated with depression compared to genetic factors associated with educational attainment. Given that smaller hippocampal volumes have also been associated with other psychiatric disorders, future studies should examine whether these associations are specific to depression or similar for polygenic scores for other psychiatric disorders.

### Parental History of Anxiety/Depression and PGS-DEP

In this sample of children and adolescents, 29% had a parent with a history of anxiety/depression, which is very similar to the percentage in the Adolescent Brain and Cognitive Development (ABCD) study sample (Pagliaccio et al., 2020). Having a parent with a history of anxiety/depression was associated with higher PGS-DEP in children and adolescents, consistent with previous work (Mars et al., 2022). The effect size was small, which aligns with previous research indicating that polygenic risk scores and family history measures represent related but different information (Agerbo et al., 2021; Loughnan et al., 2022; Lu et al., 2018). There was an indirect association between parental history of anxiety/depression and anterior hippocampal volume via PGS-DEP. Having a parent with a history of anxiety/depression was significantly associated with higher PGS-DEP, which was in turn associated with smaller anterior hippocampal volume. These findings suggest that one pathway through which family history of depression may lead to reductions in anterior hippocampal volume for children is through increases in genetic risk. Given that parental depression is also associated with more stressful rearing environments for children, future studies should disentangle how much the transmission of risk across generations, including at the neural level, is due to genetic and environmental influences (Goodman & Gotlib, 1999).

Parental history of anxiety/depression was not directly associated with anterior or posterior hippocampal volume in children and adolescents. While some previous studies have shown associations between parental history of depression and hippocampal volume in children or adolescents (Chen et al., 2010; Rao et al., 2010), others have not (Lupien et al., 2011; Mannie et al., 2014; Pagliaccio et al., 2020). These findings may be partially attributable to the type of family history assessment used. A more rigorous, clinical interview measure of parental history of depression may have yielded significant results.

### Strengths and Limitations

Key strengths of this MRI study include the relatively large sample size coupled with the use of PGS-DEP derived from one of the largest recent GWAS of depression (Howard et al., 2019). Another major strength of this study stems from the careful implementation of a combined automated and manual segmentation approach to computing hippocampal subregional volumes (Decker et al., 2020). At the same time, several limitations of this study must be considered when interpreting the results. First, this study utilized a cross-sectional and correlational design. Second, analyses of PGS-DEP and PGS-EA included only participants of European ancestry, similar to many studies involving polygenic scores (Elliott et al., 2019; Kwong et al., 2021; von Stumm et al., 2020). Therefore, the findings from this study are not generalizable to populations beyond individuals of European ancestry. Third, data on depression symptoms were only collected for a small subsample, precluding analyses of the role of depression symptoms.

## Conclusion

Findings from this study link higher polygenic risk for depression with reduced anterior but not posterior hippocampal volume in children and adolescents. Higher genetic risk for depression may lead to reductions in anterior hippocampal volume that contribute to the onset or worsening of depression. Parental history of depression may lead to smaller anterior hippocampal volumes in children in part by increasing children’s genetic risk for depression. These findings can be used to inform the design of more effective prevention and intervention strategies for depression. Anterior hippocampal volume could be investigated as a biomarker to match individuals with effective treatments. Early identification of children at risk for depression can be facilitated by increased knowledge of genetic and neural markers measurable prior to the onset of depression or early in the course of the disorder.

## Data Availability

This study utilized data from the Pediatric Imaging, Neurocognition, and Genetics (PING) study, which is publicly available (https://nda.nih.gov).

https://www.nitrc.org/projects/ping/

