## Supplementary Material for "Polygenic Risk for Depression and Anterior and Posterior Hippocampal Volume in Children and Adolescents"

### Supplemental Material

#### Adjusting for Intracranial Volume

As described in Decker et al. (2020), a regression approach was used to correct regional volumes for intracranial volume (ICV) (Jack et al., 1989). In this analysis, intracranial volumes were regressed onto regional hippocampal volumes (left and right, anterior and posterior), such that the residual value (the regions size minus its predicted value based on each individual's ICV) was accounted for in each region for each individual. Before correcting for ICV, four separate linear regression models were fit to test whether age or sex interacted with ICV to predict left and right anterior and posterior hippocampal volumes. While sex did not interact with age or ICV to predict volumes, the interaction between ICV and age was significant for left and right anterior hippocampal volumes. Therefore, the sample was divided into subsamples based on age for the anterior hippocampus, and ICV correction was performed separately on these subsamples. In adjusting the volume of the right and left anterior hippocampus, younger and older children (3–7, 8–12 years of age) were combined into a single group because the association between ICV and volumes did not differ in younger and older children. For the same reason, adolescents and young adults (13–17, 18–21 years of age) were combined into a different group for ICV correction. Since neither age nor sex interacted with ICV to predict posterior hippocampal volumes, the sample was not divided into subsamples to adjust posterior hippocampal volumes for ICV. After running further analyses described in Decker et al. (2020), bilateral hippocampal volumes were calculated by summing analogous regions in the left and right hemispheres. Bilateral hippocampal volumes that had been adjusted for ICV were used in all analyses.

**Table S1.** The number of SNPs from the Howard et al. (2019) GWAS included in each PGS-DEP *p*-value threshold

| **PGS-DEP *p*-value threshold** | **Number of SNPs from Howard et al. (2019) included** |
|---|---|
| 1 x $10^{-6}$ | 161 |
| 1 x $10^{-5}$ | 341 |
| .0001 | 811 |
| .001 | 2400 |
| .01 | 8438 |
| .05 | 20955 |
| .1 | 31363 |
| .5 | 75796 |
| 1 | 97276 |

*Note*. SNP, single nucleotide polymorphism; GWAS, genome-wide association study; PGS-DEP, polygenic risk score for depression

**Table S2.** Zero-order correlations between PGS-DEP, parental history of anxiety/depression, and anterior and posterior hippocampal volume

| | | 1 | 2 | 3 | 4 | 5 | 6 | 7 | 8 | 9 | 10 | 11 | 12 |
|---|---|---|---|---|---|---|---|---|---|---|---|---|---|
| 1 | PGS-DEP at $p < 1$ | -- | | | | | | | | | | | |
| 2 | PGS-DEP at $p < .5$ | .996*** | -- | | | | | | | | | | |
| 3 | PGS-DEP at $p < .1$ | .92*** | .93*** | -- | | | | | | | | | |
| 4 | PGS-DEP at $p < .05$ | .88*** | .88*** | .95*** | -- | | | | | | | | |
| 5 | PGS-DEP at $p < .01$ | .75*** | .75*** | .81*** | .86*** | -- | | | | | | | |
| 6 | PGS-DEP at $p < .001$ | .58*** | .58*** | .63*** | .65*** | .78*** | -- | | | | | | |
| 7 | PGS-DEP at $p < .0001$ | .44*** | .44*** | .48*** | .49*** | .60*** | .80*** | -- | | | | | |
| 8 | PGS-DEP at $p < 1 \times 10^{-5}$ | .32*** | .32*** | .36*** | .38*** | .49*** | .65*** | .81*** | -- | | | | |
| 9 | PGS-DEP at $p < 1 \times 10^{-6}$ | .26*** | .26*** | .30*** | .33*** | .41*** | .55*** | .69*** | .88*** | -- | | | |
| 10 | Parental history of anxiety/depression | .09$^{+}$ | .08$^{+}$ | .10* | .09* | .06 | .07 | .04 | .05 | .05 | -- | | |
| 11 | Anterior hippocampal volume | -.08 | -.08 | -.11* | -.11* | -.08 | -.12* | -.05 | -.07 | -.08 | -.03 | -- | |
| 12 | Posterior hippocampal volume | .05 | .04 | .02 | .01 | -.02 | -.02 | -.06 | -.07 | -.07 | .001 | .07* | -- |

*Note*. *n* for correlations between PGS-DEP and parental history of anxiety/depression = 522; *n* for correlations between PGS-DEP and hippocampal volume = 350; *n* for correlations between parental history of anxiety/depression and hippocampal volume = 703.

$^{+}$ $p < .10$, * $p < .05$, ** $p < .01$, *** $p < .001$

**Table S3.** Associations between PGS-EA and anterior and posterior hippocampal volume

| | Anterior hippocampal volume | | | Posterior hippocampal volume | |
|---|---|---|---|---|---|
| PGS-EA *p*-value threshold | *β* | *p* | | *β* | *p* |
| 1 | .06 | .2651 | | -.07 | .2197 |
| .5 | .07 | .2172 | | -.06 | .3079 |
| .1 | .06 | .2831 | | -.06 | .3036 |
| .05 | .05 | .3524 | | -.06 | .3017 |
| .01 | .04 | .4271 | | -.05 | .3195 |
| .001 | .02 | .7325 | | -.02 | .6736 |
| .0001 | .04 | .4475 | | .02 | .6919 |
| $1 \times 10^{-5}$ | .06 | .2360 | | .03 | .5359 |
| $1 \times 10^{-6}$ | .04 | .4826 | | .07 | .2106 |

*Note*. PGS-EA, polygenic score for educational attainment

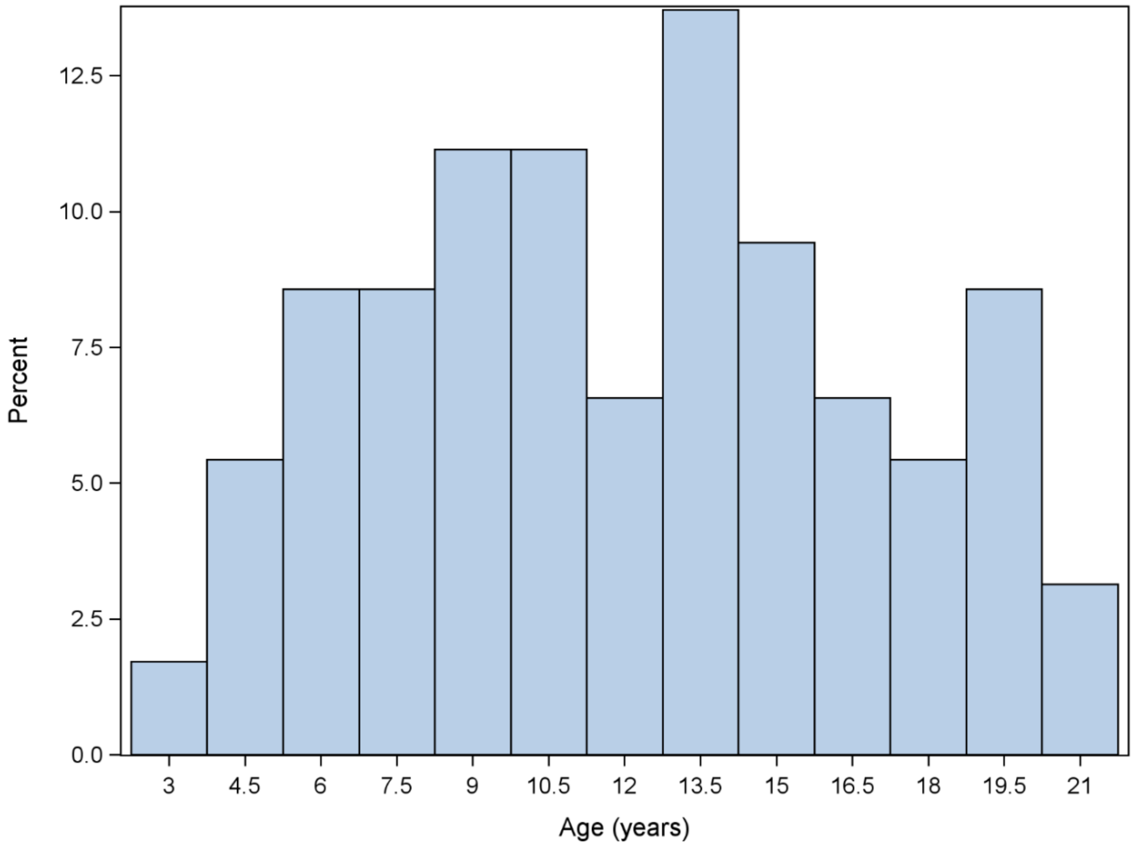


**Figure S1.** Histogram showing the age distribution ($N$ = 350)
